# EpiPathAI: Using Large Language Models to Explore Mechanisms of Life Course Exposure-Outcome Associations

**DOI:** 10.1101/2024.10.17.24315648

**Authors:** Shuang Wang, Yang Zhang, Darui Gao, Ying Gao, Xin He, Guanghui Deng, Wuxiang Xie, Jian Du

## Abstract

Large language models (LLMs) enhanced with Graph Retrieval-Augmented Generation (GRAG) are promising for life-course epidemiology, which typically depends on costly and incomplete cohort data. Inspired by the epidemiological pathway model, we introduce EpiPathAI, which combines literature-derived causal knowledge graphs with LLMs to mine bridging variables and synthesize potential mechanisms between gestational diabetes and dementia. We test four GRAG strategies on GPT-4 and evaluate the identified mediators with clinical experts and three other LLM reviewers. The knowledge graph identifies 118 bridging variables, including coronary heart disease and chronic kidney disease, previously validated in our data-driven approach through the UK Biobank. EpiPathAI has identified additional clinically meaningful mediators, including high-level low-density lipoprotein (9.8% of effect, 95% CI: 3.7%-23.2%), and depression, which is a reasonable but statistically non-significant mediator in UK Biobank. EpiPathAI serves as a knowledge-driven mechanism mining agent that complements the data-driven approach, providing a compelling foundation for investigating other mediating pathways in future longitudinal cohort studies.

## Introduction

Understanding the complexity of mechanisms and establishing causality across the life course is crucial to developing preventive and management strategies for chronic diseases [1]. Conceiving life as continuums, causal effects research across lifespan aims to study how physical and social exposures throughout all life stages affect health outcomes in later life, either individually, cumulatively, or interactively [2, 3]. This allows the development of early intervention in the modifiable factors that contribute to adverse outcomes [4]. However, life course epidemiology research faces methodological and practical challenges. Instead of case-control studies or clinical trials [5], this field relies heavily on longitudinal cohort studies, which are costly, time-intensive, and require long-term follow-ups, limiting their reproducibility and generalizability. Can findings from a generation born 50 years ago be applied to the current generation? Additionally, measuring exposures across the life course is a major source of bias, often relying on incomplete or poor-quality data [1]. Thus, it is essential to design a reliable, efficient, and generalizable automated system to explore the potential mechanisms and trajectories of a given pair of exposure and adverse outcome in life cycle.

As biomedical literature contains extensive knowledge on disease mechanisms linking risk factors to disease outcomes, literature-based discovery (LBD) provides new opportunities for mining the mechanisms and pathways bridging exposure and outcome of interest, especially in exposure-cancer associations [6–11]. LBD attempts to identify unknown knowledge implicitly in the entire literature corpus rather than explicitly stated in one article. If one article asserts that “A affects B” and another that “B affects C”, then “A affects C” is a natural hypothesis. This A-B-C model is quite similar to the pathway model (also known as chain-of-risk model) in life course epidemiology, which focuses on the sequential link between multiple exposures, namely risk A *increases* the likelihood of risk B and then risk B *contributes to* outcome C [1]. Bristol University has developed two A-B-C model-based tools, i.e., Text Mining for Mechanism Prioritisation (TeMMPo) and Mining Enriched Literature Objects to Derive Intermediates (MELODI) [12, 13], to identify the set of potential intermediate mechanisms. A two-stage approach was built to 1) identify potentially novel intermediate phenotypes (IPs) through LBD and 2) systematically review the associations of exposure-IP and IP-cancer. However, both the non-synthesized trivial variables (a set of IPs) from stage 1 and the labor-intensive work of systematic review in stage 2 limit its scalability and generalizability. Given the massive intermediate candidates, the essence of LBD is to rank the important pattern “A affects C via B” that is novel, plausible, and clinically meaningful. Such information could be used to enhance our understanding of the causality behind identified associations and inform the planning of further investigations [14–16].

Large language models (LLMs), with excellent capacities for summarizing large-scale text, have the potential to synthesize piecemeal findings and reason the risk chain linking the exposure to outcome [17]. However, LLMs may generate inaccurate content with hallucinations [18]. Retrieval Augmented Generation (RAG) is a cost-effective strategy by retrieving the most relevant and up-to-date external sources for more accurate responses [19]. However, naïve RAG and its variations focus on extending the context window to allow for longer external texts, ignoring the graphic structure of semantic concepts within scientific texts, resulting in limited performance in complex reasoning questions, such as potential chain of risk and mechanisms of exposure and outcome [20]. Graph RAG (GRAG) provides promising solutions to alleviate the problem. A well-established and fact-based knowledge graph (KG) can be a great graph index locating relevant information of query terms, through graph topological metrics, sub-graph identification [21], and community detection [20]. Based on the graph analysis, one may locate more relevant contexts feeding LLMs to generate more completed and accurate underlying information for intermediating the observed life-course correlations from cohort data.

Dementia, a typical later-life condition, is the world’s seventh-leading cause of death [22] and women are suffering from more impacts by dementia [23]. In our recent previous cohort study, we [24] found that women with a history of gestational diabetes mellitus (GDM) are linked to a higher risk of dementia in later life, first revealing the connection between these two pivotal phrases in life. GDM, as the most common (prevalence ranging from 11.7-30.2%) pregnancy complication [25], associated with various maternal and fetal adverse outcomes during pregnancy, plays a crucial role in lifespan and also increases the risk of long-term metabolic and cardiovascular diseases in women and their offspring. However, currently, scarce research has studied the relationship between GDM and dementia, let alone explored the underlying mechanisms. Considering the rare effective treatment of dementia and the great dementia-based burden on women (higher mortality and dementia-based disability-adjusted life years) [23], it is imperative to provide scalable and reliable automated solutions to delineate the trajectories and potential mechanisms linking GDM to dementia. This will facilitate the identification and early intervention of the modifiable factors before aging in GDM mothers.

Inspired by the pathway model in life course epidemiology [1], integrating the idea of LBD, and LLMs’ powerful summarization and reasoning capacity, we present a novel automatic system ‘EpiPathAI’, to map the risk trajectory from GDM to dementia and other neurodegenerative diseases (abbreviated as dementia in this study) (Fig 1). Here we utilized 35,010 semantic triples (subject-predicate-object) from SemMedDB [26] extracted from 28,280 higher argument strength sentences within 14,733 GDM and Dementia articles published in high-quality journals. This dataset was used to construct a causal knowledge graph related to GDM and Dementia (GDM-Dementia). In particular, we fine-tuned Llama2-7b [27] to identify sentences with stronger argument strength which outperforms the GPT-3.5 turbo (few-shot). We then identified the core bridging variables through network mining techniques and conducted evaluation and prioritization with the GPT-4 [28] assessment and human fact-check. Further, with GPT-4 as the baseline model, we compared it with three other RAG strategies derived from the above gradual network mining methods to summarize and explore the potential GDM-Dementia mechanisms. Evaluations of those results were completed by clinical expert reviewers and three LLMs reviewers (GPT-4o [29], Llama3-70b [30], Gemini Adv [31]). Finally, based on all the above findings, we took the tentative step to discuss the potential GDM-Dementia mechanisms. Moreover, utilizing 205,463 women participants (GDM history, 1292; non-GDM history, 204,171) from the UK Biobank [32], a long-term cohort since 2006 covering various health data of over half a million people from 40-69 years, we measured the mediating effects of three representative mediator candidates explored by EpiPathAI to further validate the effectiveness of the identified variables and mechanisms.

**Fig 1.**
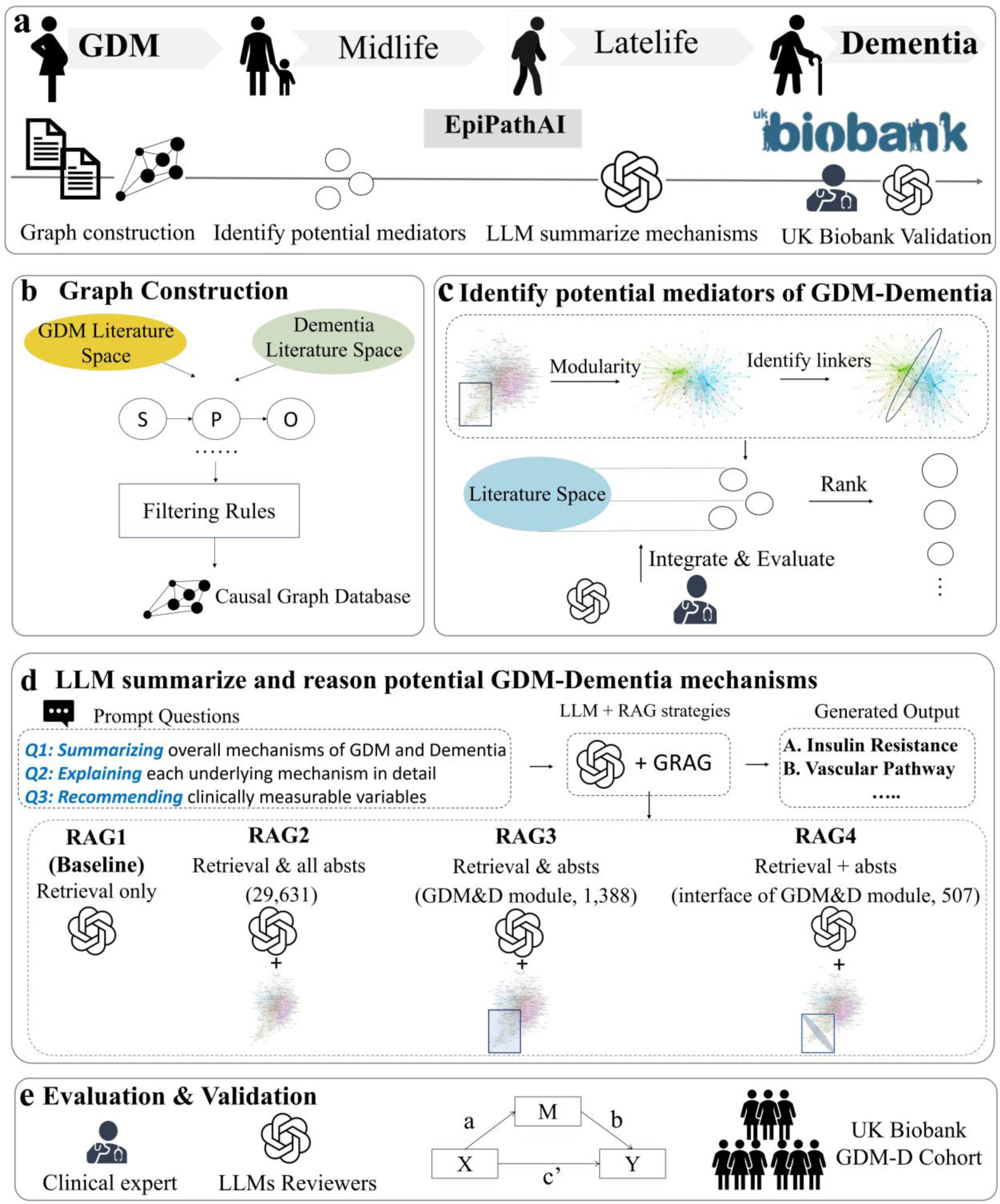
The progressive research pipeline of EpiPathAI. | **a.** The research overview of EpiPathAI. **b-e** are detailed exhibitions of its four components. In component **d**, four RAG strategies assisting GPT-4 were compared in summarizing and reasoning potential mechanisms of GDM-Dementia. GDM-D indicates GDM-Dementia. RAG1 indicates the baseline GPT-4 model using the original GPT-4 RAG retrieval technique. RAG2 indicates the basic GPT-4 attached with all abstracts of nature index journals as the external knowledge base. RAG3 and RAG4 are GRAG strategies. RAG3 uses the basic GPT-4 with all abstracts in the sub-community of GDM and Dementia. RAG4 uses the basic GPT-4 with refined abstracts of the top quantile 50 intermediating variables of the subcommunity of GDM and Dementia. In section **e**, LLMs reviewers include three models, GPT-4o, Llama3-70b, and Gemini Adv.

## Results

### Descriptive analysis of the pruned causal GDM-Dementia KG

Firstly, Fig 2 shows the overall process from building to pruning a causal GDM-Dementia KG using strong argumentation contexts, including filtering aspects of journal type, edge (predicate) type, node (node) type, and argument strength of semantic triple’s source context. In classifying the argumentation strength of context sentences, we fine-tuned Llama2-7b with a manual annotation set of 576 sentences, which exhibits a better performance (accuracy, 0.84; F1 score, 0.70) than the GPT-3.5 turbo few-shot (Supplementary Table 2). We then applied our fine-tuned Llama2-7b to 61,331 source sentences for the cogency classification task. Among them, 52,347 (85.35%) sentences were classified as strong argument strength, either prior knowledge, new findings, or recommendation on new findings, and 8,984 weak argumentation sentences (i.e., research hypothesis, research design statements) were excluded. See methods for more details on the KG built and argumentation classification process.

**Fig 2.**
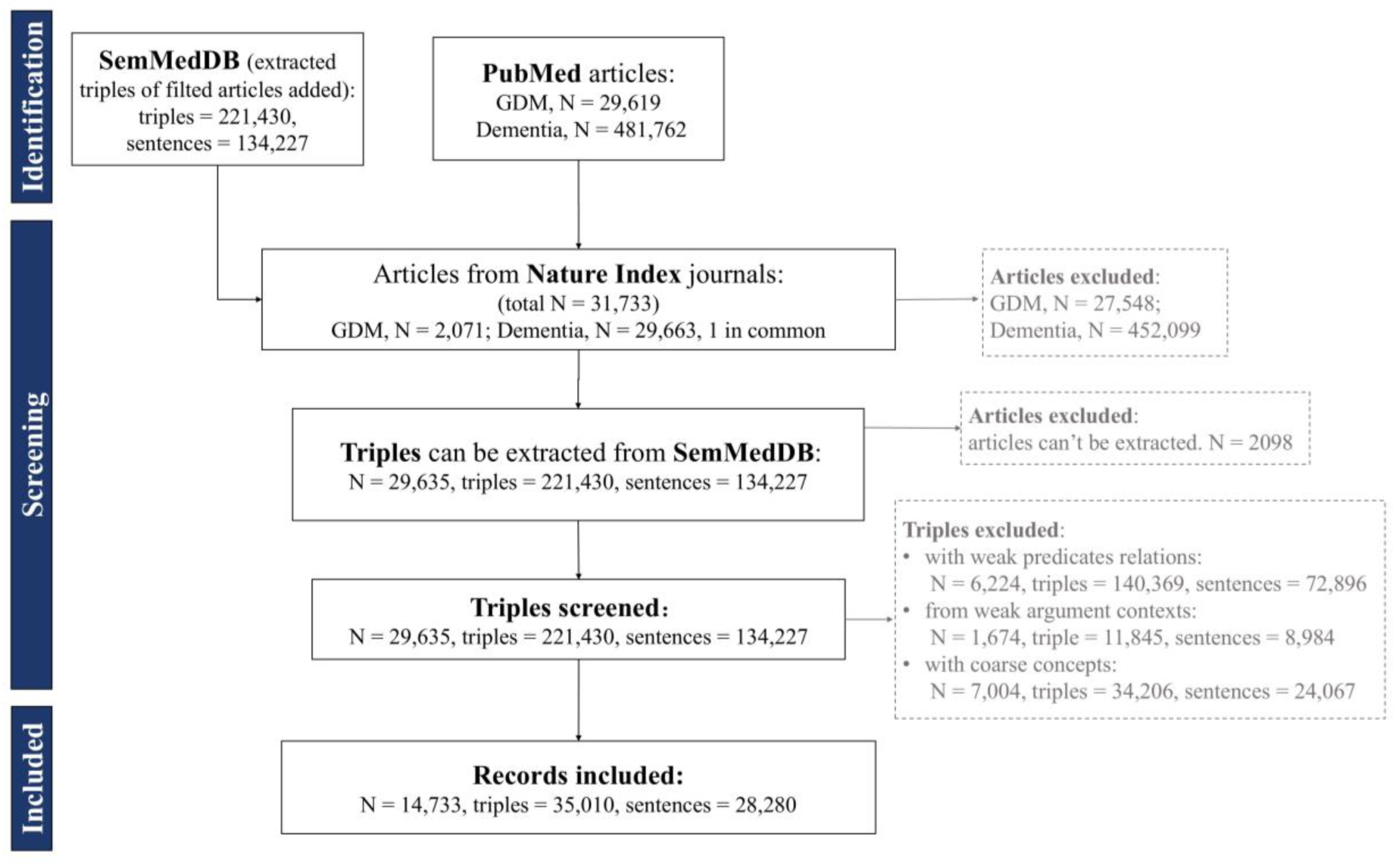
The PRISMA workflow chart of data collection and processing process of building and pruning the GDM-Dementia KG. N indicates the count of papers. Dementia articles include all articles relevant to Dementia and other neurodegenerative diseases.

After further screening semantic triples, a total of 8,488 unique concepts and 35,010 relations were finally included for research, from 28,280 unique context sentences in abstracts of 14,733 articles. Among them, the leading three concept types are Genes & Molecular Sequences (36.97%), Chemicals & Drugs (36.46%), and Disorders (16.32%) and the leading three relations are ASSOCIATED_WITH (21.77%), AFFECTS (19.61%), and CAUSES (16.92%).

Please refer to Supplementary Table 3-4 for the overall distribution of concept types and relations types.

### Identification and evaluation of key bridging nodes

Next, we conducted community detection using the Louvain modularity algorithm to detect the network structure of the GDM-Dementia KG. With each pair of concepts as nodes and the count of predicate as their edge weight, the initial graph contains 8,488 nodes and 23,214 edges. As a result, 8,488 unique nodes were partitioned into 228 communities (Fig 3a). Interestingly, we found that ‘Gestational Diabetes’ and ‘Dementia’ were allocated into the same community, indicating that they are connected tightly.

**Fig 3.**
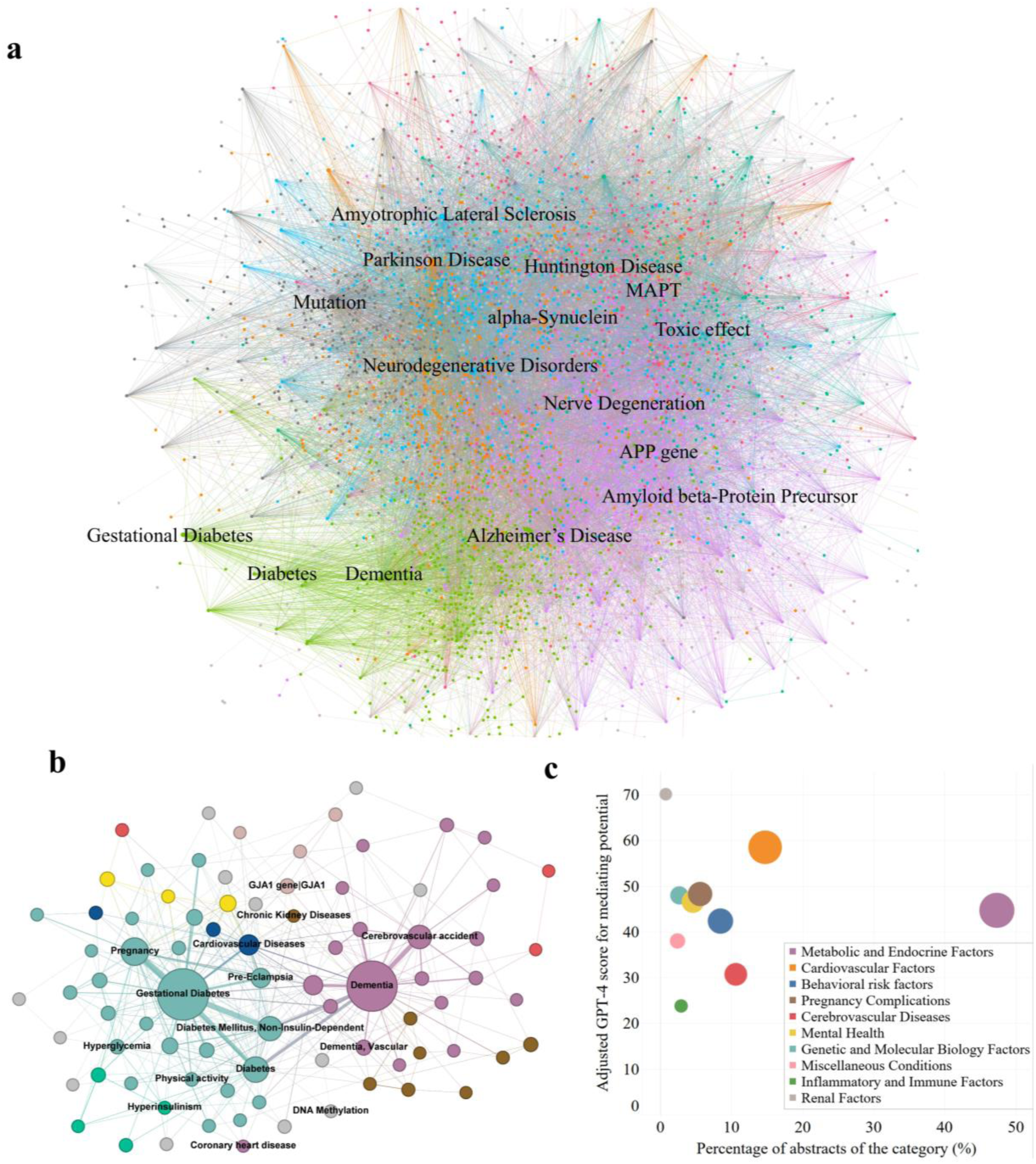
The modularity mining of the GDM-Dementia network and the rank of core bridging variables |. **a**. The network modularity of the GDM-Dementia network. The top 54.91% of degrees’ nodes (N = 170, degree >= 2) were chosen for optimal visualization. **b.** The sub-network modularity of the community involving GDM and Dementia. The graph exhibits the top 8.89% degrees’ nodes (N = 82, degree >= 7) for optimal visualization. The bigger the node, the higher the weighted degree of the node. **c**. The mediating potential and reliability of each category from GDM to Dementia. The x-axis exhibits the percentage of abstracts of all variables of each category in all abstracts of 118 leading mediating candidates. The y-axis shows the average adjusted GPT-4 evaluation scores of representative variables in each mediator category, indicating the reliability of mediating potential.

To further dig into the connections between GDM and dementia, we narrowed down the mining zone to the sub-community (924 nodes and 1535 edges) involving both GDM and dementia. We re-analyzed this sub-community and detected 24 communities inside it centered on GDM and dementia (Fig 3b). Surprisingly, among all bridging nodes (N = 235), type 2 diabetes, coronary heart diseases, chronic kidney diseases, physical activity, and comorbidity were validated as mediators for women with a history of GDM to develop long-term dementia by Zhang et al [24]. This study was not included in our KG, serving as an ideal outer validation to prove the tentative effectiveness of our intermediating mining approach. Also, the 2024 report of the Lancet Commission on dementia prevention, intervention, and care, identified 14 potentially modifiable risk factors being adopted to policy and clinical guidelines, all factors except low educational attainment in early life were identified by our bridging candidates [33]. Inspired by these great correspondences, we believe those pivotal bridging nodes may have great potential as GDM-Dementia mediators. The final filtered core bridging mediator set included 118 variable candidates consisting of the top quantile 50 of all bridging nodes.

After GPT-4 automatic categorization and human review, 108 mediator candidates were left and categorized into three classes in level 1 and ten sub-classes in level 2 for further class-based potential mechanism prioritization (Supplementary Table 5 for all variables). For representative variables of each class, feeding on all abstracts indicating GDM-X and X-Dementia in KG, GPT-4 was utilized to review and rate the mediating potential of the chosen mediator, and further human evaluation on GPT-4 reviews were conducted. The blind-evaluation results of two students revealed that, though fact hallucination and argumentation deficits exist in GPT-4 review results, the overall GPT-4 evaluation quality is fundamentally reasonable, with an average of 6.85 in factuality (consistency of evidence fact and review results) and 6.40 in argumentative logic rationality (Supplementary Table 6). Combining GPT-4 review scores and blind-evaluation scores of those review scores as the adjusted GPT-4 review scores showed that the top three mediating categories are renal factors, cardiovascular factors, and pregnancy complications (Supplementary Fig 1). Out of expectation, the biggest and most well-known mediating category of metabolic and endocrine factors (31 factors supported by 196 abstracts) dropped out of the top three. This might be because, though not exceeding the token limit, with the big volume of texts in place, GPT-4 is inclined to be ‘lazy’ and only read partial abstracts for a generation. Also, when it comes to more common knowledge (i.e., the association of obesity and GDM), it tends to mismatch the article index to its already known information instead of the article’s real content, lowering its factuality scores in human evaluation.

Due to the above concerns, we further integrated the literature volume of each category and adjusted GPT-4 scores to collectively visualize the mediating potential of each mechanism category (Fig 3c). We found that the metabolic and cardiovascular factors are on the top right with relatively large evidence volumes and high mediating scores. Worthy to note, though renal factors are supported by merely three abstracts in the filtered KG, the assertive claims of the elevated risk chronic kidney disease (CKD) after GDM irrespective of the subsequent development of diabetes [34] and the independent risk factors of CKD and acute kidney injury (AKI) on central nervous systems [35] gave GPT-4 more assurance for a high mediating score on renal factors.

### LLM synthesis and reasoning on potential mechanisms and actionable variables

Besides variable level mining of underlying mechanisms through graph mining, we also explore and compare the above graph mining-based RAG strategies (RAG1: the baseline GPT-4 model (retrieval-only); RAG2: the baseline GPT-4 + all nature index (NI) journals’ abstracts; RAG3: the baseline GPT-4 + all NI abstracts of the GDM-Dementia sub-community; RAG4: the baseline GPT-4 + all NI abstracts of the Q50 bridging variables of the GDM-Dementia sub-community, Fig 1c) in assisting LLMs with potential mechanisms summarization and reasoning. Specifically, we designed three questions: 1) summarize and reason the overall underlying mechanisms of GDM-Dementia, 2) conduct deep mining of each underlying mechanism, and 3) draft a plan (indicators to measure) for constructing a cohort of GDM women for the prevention of long-term dementia (Appendix A of supplementary file for the detailed questions). For each RAG strategy, we utilize GPT-4 to answer all three questions and have clinical experts (grading results see supplementary table 7-8) and three LLM reviewers (GPT-4o, Llama3-70b, Gemini Adv) rate each answer from three dimensions, scientific accuracy & completeness, novelty, and clinical relevance (only for Q3). In general, we found that for each metric and question, review grading scores from all reviewers incrementally increased from the baseline GPT-4 (RAG1) to the RAG with refined bridging variables’ contexts (RAG4) (Fig 4). This primarily reveals that from the naïve RAG baseline to the refined GRAG strategy, gradually optimized RAG strategies are beneficial in improving the quality of GPT-4 answers.

**Fig 4.**
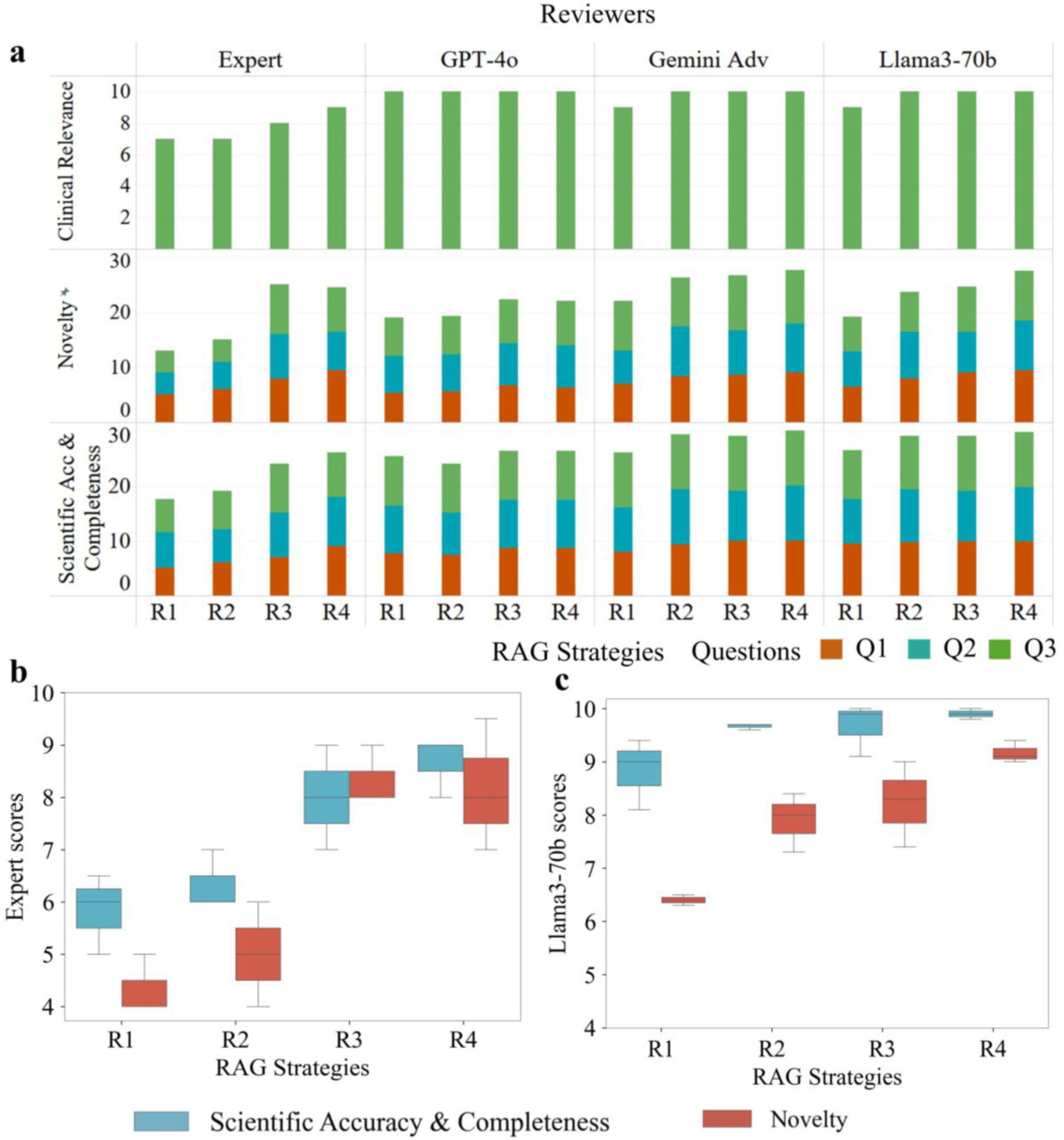
Scoring trends of different responses based on RAG 1-4 in different reviewers. | **a**. Scoring trends of different RAG-type answers in expert-reviewer and LLMs-reviewers. Scientific acc & Completeness indicates the metric of scientific accuracy & completeness. **b**. The distribution of expert scores by RAG strategy and Metric. Clinical relevance is excluded from the box chart distribution, as each RAG type only has a data point of clinical relevance applicable to Q3. **c**. The scores distribution of Llama3-70b, as a representative LLM reviewer, by Response and Metric.

Further, to figure out the statistical difference among each dimension, we conducted a single-factor ANOVA analysis of expert scores of answers across all three dimensions separately (RAG type, metric type, and question type). The results of the Homogeneity test of variance using Levene’s Test showed no significant heterogeneity in the variances of scores within each dimension (Results 3.1 of the supplementary file), allowing the subsequent ANOVA analysis. We find that RAG strategy type significantly (P < 0.05) contributes to scores’ differences, while no significance was detected regarding metric type and question type (Supplementary Table 9A). The post-hoc analysis reveals that the mean scores of outputs of RAG4 incorporating contexts of all bridging variables (8.50) and RAG3 including contexts of the GDM-Dementia subcommunity (8.14) are significantly (P <= 0.001) higher than the means of RAG2 with all unfiltered abstracts (5.86) and GPT-4 baseline (5.36). However, no significant difference was found between RAG4 and RAG3 or RAG2 and RAG1 in expert scores (Supplementary Table 9B). These results were supported by LLMs reviewers (Llam3-70b and Gemini Adv) with reviewing scores of the bridging-context RAG4 significantly higher than that of the GPT-4 baseline (Results 3.2 of the Supplementary file). This suggests that the modularity-filtering method did help target relevant knowledge and supplement pivotal information to GPT-4 to finally improve the overall quality of the generative answer regardless of question and metric types.

Moreover, we delved deeper for more details about the only significant factor of RAG strategy type. We analyzed ‘metric’ and ‘question’ separately under each stratification of RAG strategies. Surprisingly, in terms of metric type, we found that in GPT-4 baseline, the average novelty score (4.33) is significantly (P < 0.05) lower than the average of scientific accuracy & completeness (5.83) (Supplementary Table 9C). This suggests that with the baseline GPT-4 RAG, though the scientific accuracy of generation is appropriate, it is hard to capture novel and surprising information. However, other RAG strategy tiers make up this point, rendering both novel and accurate generative content with no significant differences found. In addition, no significant differences in scores of different questions are found under each RAG strategy (Supplementary Table 10).

### Detailed formulation of underlying GDM-Dementia mechanisms and UK Biobank cross-validation

Finally, after the RAG strategies comparison experiments, integrating mining results of all four versions of GPT-4 generations, we summarized and reasoned the main five potential mechanisms and intermediates involved in the association between GDM and dementia (Fig 5, original GPT-4 generations see Appendix B of the Supplementary file). A-C are quite traditional and well-known factors that explain the association between DM and dementia [35]. Considering the similar pathophysiological mechanisms between GDM and type 2 diabetes, and women with GDM history are at high risk of developing postpartum DM throughout their later years [36]. It is very reasonable to explore these mediating factors between GDM and dementia to cover the above content [36] and factors of A and B were already validated by former UKB-based work [24]. Specifically, except for type 2 diabetes, coronary heart diseases, EpiPathAI also identified bridging factors of chronic kidney diseases, comorbidity, and physical activity, validated as mediators of GDM-Dementia previously [24]. As for the other mediators explored by EpiPathAI, we then respectively tested the mediating role of three representative potential mediators: high LDL-C (lipid metabolism), depressed mood, and early menopause (hormonal changes) in the same former UKB cohort [with 205,463 women participants (GDM history, 1292; non-GDM history, 204,171)], for cross-validation. Results suggest that only high LDL-C significantly mediated the association between GDM and dementia, as shown in Figure 5-b. High LDL-C, explained 9.8% (95% CI: 3.7%-23.2%) of the observed association. The above correspondence suggests that as a knowledge-driven system, EpiPathAI results are aligned with most of the results of the UKB cohort-based data-driven method.

**Fig 5.**
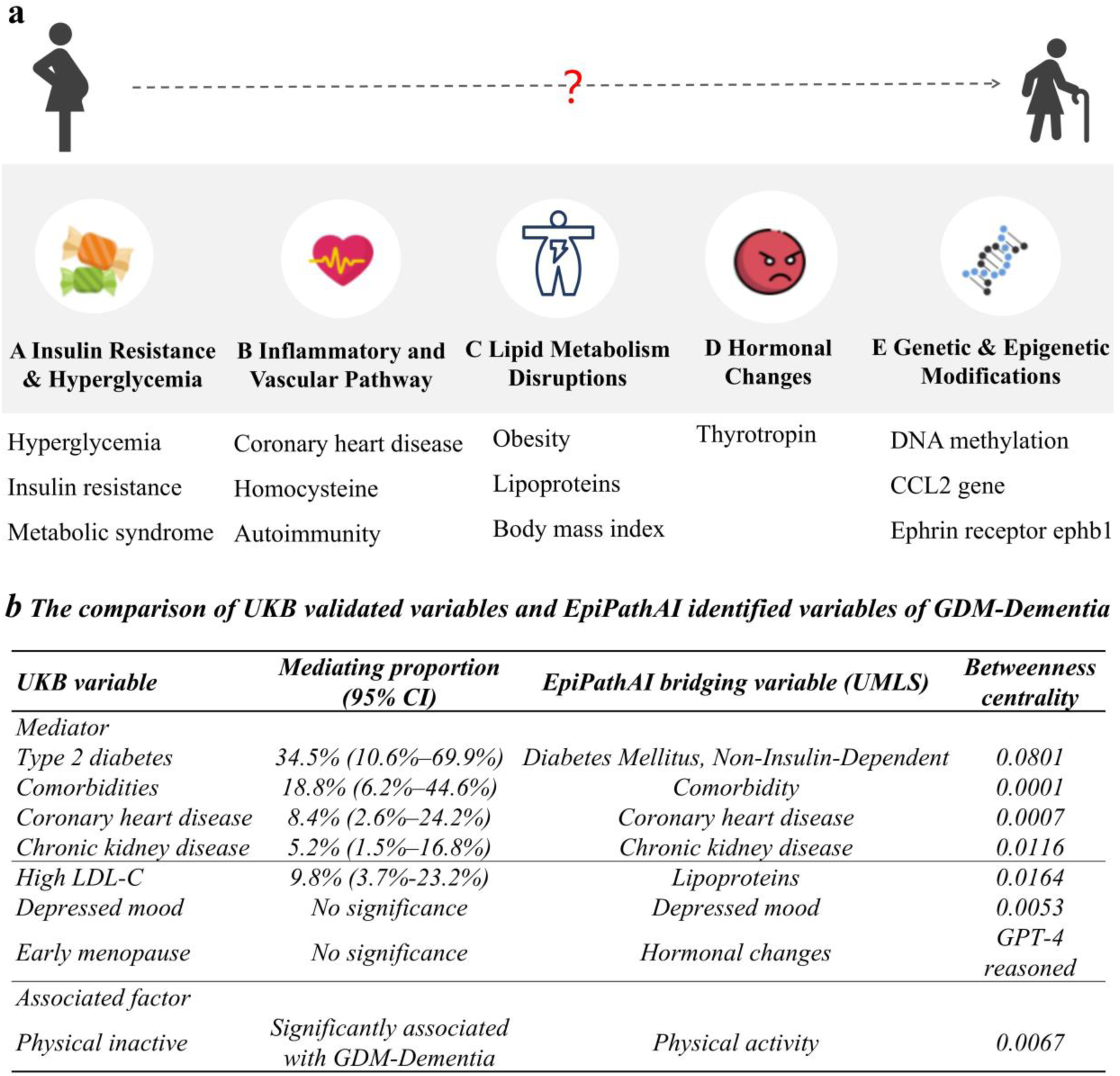
Five potential mechanisms generated by GPT-4 and UK Biobank validation. | **a.** Five potential mechanisms from GDM to Dementia summarized and reasoned by GPT-4. Below each potential mechanism, representative bridging variables in the filtered casual graph are listed. In table **b**, the mediating proportion results of type 2 diabetes, comorbidities, coronary heart disease, and chronic kidney disease and the statistical result of physical inactive referred from the former study [24]. Comorbidities (from UKB variable) were defined as the diagnosis of any two or three of the three chronic non-communicable diseases: type 2 diabetes, coronary heart disease and chronic kidney disease. Comorbidity of the UMLS term refers to the general status of the co-existence of two or more disease. Early menopause is defined as menopause age<40 years old.

Though the mediating effect of either early menopause or depressed mood was not significant based on UKB cohort, there may be multiple reasons for this, such as the limitations of using only early menopause to represent hormone effects. EpiPathAI provides great insights in the potential mechanisms of GDM-Dementia beyond. Instead of a specialized GDM long-term cohort, UKB has only limited cases of observed conditions, EpiPathAI should not be confined within it, but rather extend beyond it. Multiple hormonal changes during and after pregnancy, such as fluctuations in insulin, cortisol, estrogen, and progesterone levels, all play direct or indirect roles in modulating A and B and contribute to dementia [37]. Furthermore, the impact of estrogens extends beyond the reproductive system to various brain functions, including learning, memory, and neuroprotection against dementia [38]. From a long-term postpartum perspective, the decline in estrogen during menopause can exacerbate the risk of cognitive decline in women with a history of GDM. That is to say, the potential link between GDM and dementia may be related to multiple hormonal changes at different stages of life, encompassing both the pregnancy and postpartum periods.

The most commonly mentioned genetic and epigenetic modifications during pregnancy are changes in glucose and lipid metabolism in the uterine environment of women with GDM, which have short-term or even long-term effects on the fetus by altering the epigenetic modifications of their offspring genes [39]. However, epigenetic modification in adults can also be caused by various factors, such as stress, air pollution [40], and malnutrition [41]. Therefore, it is reasonable to believe that epigenetic changes during pregnancy due to hyperglycemia or other metabolic dysfunction may lead to lasting changes in gene expression affecting not only offspring but also GDM women themselves.

Based on the intermediary factors mined by EpiPathAI, we fully recognized that hyperglycemia, insulin resistance, dyslipidemia, inflammatory response, vascular change, genetic and epigenetic modifications, and hormone changes are all involved in the pathogenesis of GDM and dementia, which not only limited during pregnancy but may still exist after childbirth or reappear after a period of time, contributing to the development of GDM into long-term dementia. Furthermore, the aforementioned pathways do not exist independently but will influence and promote each other, linking the seemingly unrelated diseases of GDM and dementia together.

## Discussion

This study proposes a novel EpiPathAI system to explore mechanisms linking life-course exposures and outcomes when lacking high-quality cohort data. Integrating GDM and Dementia relevant relationships reported in high argument-strength statements of scientific literature, we identified key bridging variables in the interface of GDM and Dementia using graph community detection and centrality metrics, including factors such as type 2 diabetes and chronic kidney diseases which were validated as mediators priorly. To improve the understanding and interpretability of GDM-Dementia mechanisms, we fully leveraged the advantages of GPT-4 in evidence synthesis and reasoning and compared four different RAG strategies to identify novel and reliable mechanisms and clinical modifiable indicators. After expert evaluation, results showed that GRAG-based (sub-community abstracts; bridging variables’ source abstracts) mechanism generations are significantly superior to answers from all unfiltered articles or the GPT-4 baseline. Further, in the GPT-4 baseline, the novelty of the answer is significantly inferior to aspects of completeness & accuracy, or clinical relevance. Clinically modifiable variables can be effectively identified on the path of GDM-Dementia. This provides evidence for what patient information needs to be collected when constructing a GDM cohort and offers insights into the development of preventive strategies for dementia in the later life of GDM women.

Results indicated that conducting RAG via community detection of a refined KG significantly outperforms the naive RAG strategy in mining potential mechanisms of two given diseases, leading to more comprehensive, accurate, and novel generative content. This corresponds to the findings of Edge et al. from Microsoft, who found that community detection-based GRAG significantly exceeds the RAG baseline in answering query-focused summarization (QFS, Q&A regarding the entire private texts) questions of a private dataset [20]. In another GRAG study, Hu et al. conducted GRAG by identifying subgraphs revealing that it is significantly superior to current RAG means in complex practices needing multi-hop reasoning on textual graphs [21]. When connecting graphs and LLMs, Pan et al. present three types of interaction between knowledge graph (KG) and LLMs, “LLM-augmented KG, KG-enhanced LLM, and synergized KG and LLM” [42]. Representative studies include utilizing LLMs to create and complete KGs [43] or extracting causal graphs from texts [44]. EpiPathAI integrates these three types forming an iterative GRAG process from generating causal graphs assisted by LLMs, KG for RAG, to synthesized summarization with KG and LLM. In particular, we observed that the naïve RAG strategy is significantly worse in creating novel responses. This makes sense as the baseline RAG is limited in mining the most comprehensive, relevant, and detailed context information compared to other strategies, and thus reaches a more common and mediocre generation.

EpiPathAI has various applications for biomedical and interdisciplinary professionals and future research. First, theoretically, the present study is the first methodological and applied research regarding GRAG in the scenario of biomedical mechanisms mining. Prior studies either focus on the generic framework of medical GRAG [45] or naïve RAG research for medical guidelines on clinical disease diagnosis [46]. Starting from the specific clinical question and needs, we explored a GRAG-based streamline and validated it with the collaborative clinical expert. Second, our GRAG-based knowledge mining method can be scalable to other life cycle exposure-outcome mining issues, deepening the understanding of the *causality of life course epidemiology*. Though explicit causal inference is necessary for guiding prevention, one major issue in epidemiology is the enduring confusion between association and causality. This issue is especially true in life course, social, and environmental epidemiology in which evidence stems largely from observations and rarely from experiments such as randomized trials. It is hard to figure out the real causality fundamentally. However, our method can help delve into more details of the potential causality and be one step closer to more scientific decision-making. Moreover, these pivotal intermediate factors are potential research targets for further animal model-based experiments or other in vitro experiments, making our system a hypothesis generator to investigate the mechanisms underlying causal relationships. After verifying its scientific validity, it will also provide a theoretical basis for exploring intervention targets in the future. All these make it an effective tool for integrating clinical, basic, and translational research in life course epidemiology.

EpiPathAI has unique strengths differing from various exposure-outcome research tools in place. Compared to the Global Burden of Disease (GBD) Collaborators, which also provides the relationship between 88 exposure (i.e., risk factors) and selected health outcomes [47], though respective strengths and weaknesses exist, we believe our approach has unique advantages. Firstly, GBD Collaborators focuses on the linear relationship between individual risk factors and outcomes; whereas we explore the complex network relationships among multiple individual risk factors and outcomes. There are also interactions within risk factors or adverse outcomes. Secondly, GBD Collaborators provides the *strength of risk* associated with individual risk factors and outcomes, such as disability-adjusted life years (DALYs); we cannot provide this strength, but we offer the *strength of argumentation* by fine-tuning Llama2-7b. Third, our approach is less labor-intensive since we automatically curated a high-quality causal knowledge base related to the interested exposure-outcome pairs. Further, compared to other literature-based discovery tools, such as TeMMPo and Melodi which mainly identify mediators from the co-occurrence mindset or the flat A-B-C model (high-frequency overlapping B), EpiPathAI set out from the textual network structure through community detection and topological structure metrics to identify mediators from a global view. Also, our combination of LLMs introduces synthesized insights allowing further summarization and synthesis rather than only identifying those high-frequency overlapping concepts. This in turn makes it possible to synthesize from various evidence rather than with only the same research design evidence (i.e., only randomized controlled trials) as what TeMMPo stage 2 did.

Limitations exist in this study. First, we only included high-quality journals in the nature index list to control the research quality. Though relatively complete and promising results were revealed, in the future study, we would include all PubMed studies related to GDM and dementia, which may introduce new novelty findings. Second, GDM is a special condition impacting both mothers and fetal. The exploration of GDM women and the development of long-term dementia may be confounded by research that focuses on the association of offspring exposed to intrauterine GDM and dementia in late life. Since the extracted triple (entity-predicate-entity) is a flat relation without the attribute of the research object (mother or offspring), the initial GRAG results may involve noising mediating nodes. However, we have underlined the research objects as pregnant women in the prompt during LLMs assessment, which would control the noising effects largely. Third, candidate intermediating variables contained confounders (common cause). Though we have conducted a series of experiments of prompt engineering and designed a detailed prompt to have GPT-4 distinguish intermediating variables and confounding variables, it is still relatively difficult for LLMs to identify both accurately. Especially in medicine, many factors are bidirectionally related or the relation is contradicting and the identification itself is a domain challenge. Fourth, the token limit of GPT-4 is 128k, which would truncate extra texts in RAG strategy one when all 14,733 abstracts serve as context information. However, this truly reflected the utilization status quo of LLMs. Though models with larger token limits rapidly launch on the market, compared to the exponentially growing knowledge base volume, the key to achieving a more accurate generation is the development of proper and delicate RAG strategies.

In conclusion, integrating large-scale scientific literature, graph mining, and LLMs, this study developed and validated a systematic framework EpiPathAI to identify, synthesize, and rank novel, reliable, and clinically modifiable bridging variables and explainable mechanisms of exposure and outcome. EpiPathAI serves as a knowledge-driven mechanism mining agent that complements data-driven approach, allowing for mutual validation and the discovery of new knowledge that awaits verification. It is promising to enhance the understanding of complex life-course exposure-outcome causality, providing an evidence-based foundation for constructing high-quality long-term cohorts, thereby collectively supporting the study of life-course epidemiology.

## Methodology

We conducted the study in three steps. First, constructing a refined causal GDM-Dementia KG with filtered semantic triples of SemMedDB; Second, identifying the core bridging nodes through graph mining techniques and conducting evaluation via GPT-4 assessment and manual factuality checking; Third, based on GPT-4, comparing four RAG strategies derived from the above gradual network mining means to summarize and reason intermediating mechanisms and actionable variables of GDM-Dementia. Intermediating mechanism summarizations were evaluated by clinical expert reviewers and three LLM reviewers (GPT-4o, Llama3-70b, Gemini Adv). PubMed metadata was extracted in batch using NCBI API edirect; Data analysis and network analysis were conducted using Python 3.8; Network visualization was completed through Gephi 0.10.1; Data visualization was mainly conducted via Tableau 2022.4; Mediating analysis was conducted by SAS.

### The construction of a high-quality causal GDM-Dementia KG

To piece up the full picture of GDM and dementia, we searched and acquired GDM and dementia literature datasets separately. As of May 11, 2024, we acquired 29,619 GDM-relevant papers and 481,762 dementia-relevant papers from PubMed API. Though slight data loss via PubMed edirect API, acquiring such huge data through web search (only 10k records for a single download) is infeasible and thus we directly use API results for the research. First, to concentrate on high-quality publications, we filtered papers from the perspective of journal quality. We included only publications from 145 high-quality journals (natural science and health science) in the Nature Index list, chosen by independent researchers based on reputation, involving research fields from cell, animal, to human [48]. As a result, only 31,733 articles were included in the research. To build a knowledge graph, we extracted all subject-predicate-object semantic triples from title and abstract texts of all eligibility articles using the Semantic MEDLINE Database (SemMedDB), a repository of extracted concepts and their relations from the entire PubMed titles and abstracts [26]. We construct a weighted network with each pair of concepts as nodes (subject-object) and the relation count of each pair of nodes as edge weight.

Second, to refine a causal graph assisting in mining underlying mechanisms, we pruned the network graph across three dimensions, edge types, reliability of relations, and node types. Here, the ‘causal’ graph indicates the KG focusing on strong functionally related relations for mechanisms mining. As for edges, to keep strong semantic relations associated with biological mechanisms, such as ‘*CAUSES*’, and ‘*INHIBITS*’, we include ‘*functionally_related_to*’ and ‘*association-related’* categories of relations (see methods section of the supplementary file). Regarding the reliability of relations, we automatically evaluated the argumentative strength of the source context sentence of each triple relation and chose only strong argument relations. In general, we classify the argumentative strength of statements into four categories:

1) Hypothetical statements: lack of supporting information: research purpose, hypothesis, or methodological statements,
2) Prior knowledge: evidence based on background knowledge or prior experience,
3) Empirical findings: evidence based on authors’ empirical findings,
4) Subjective recommendations: recommendations based on authors’ findings.

We removed all relations without evidence support and kept claim relations from strong argumentative statements in categories 2 to 4. Specifically, we annotated 576 sentences as a training set and fine-tuned Llama2-7b [27] for automatic classification.

As for node filtering, we removed 262 generic nodes (i.e., Disease, Severe) including the generic node list of PubMed [49] and human-curated concepts, and kept finer-grained node classes relevant to diseases progression and pathophysiological mechanisms, including ‘Disorders’, ‘Activities & Behaviors’, ‘Genes & Molecular Sequence’, ‘Phenomena’, and ‘Physiology’. Finally, after all the above filtering and graph pruning steps, the final high argument strength causal graph contains 8,488 nodes and 35,010 triples, from 28,280 strong argumentative statements in 14,733 articles from 100 top journals. The Prisma workflow of data collection and preprocessing is shown in Fig. 2.

### Identification of pivotal bridging variables

With a well-built graph in place, we then deeply mined the network structure to reveal the potential mechanisms of GDM to dementia. We hypothesized that several steps (bridging points) might mediate the progression from GDM to dementia. For a global analysis and knowledge synthesis, we considered each ‘bridging point’ as a big community compared to the node level, enabling the extraction and summarization of mechanistic knowledge at the network community level. In other words, nodes are tightly connected within each community and the path from GDM to dementia is finally achieved after going through one or more communities. Specifically, we used the efficient and high-quality Louvain modularity algorithm to partition the communities in the refined knowledge graph with a resolution of 1. Essentially, Louvain modularity aims to assign nodes of a given network to different modules by optimizing the modularity value which measures the density within modules with respect to links between modules [50]. The algorithm iteratively allocates isolated node *i* to community *C* to optimize the modularity partition until the maximum positive modularity gain is achieved. The modularity gain can be computed as below [50, 51]. In a graph sized m, *k_i,in_* indicates the sum of all edges’ weight from *i* to other nodes in *C*, *k_i_* indicates the total weight of all edges connected to *i*, Σ_*tot*_ indicates the total weights of all edges of all nodes in *C*, and *γ* indicates the resolution value.

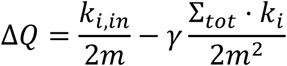

After detecting the network structure from the community level, we turn to the finer-grained level identifying bridging variables from GDM to dementia. We focus on the betweenness centrality and constraint coefficient of the structural hole. As we surprisingly found that GDM and Dementia are located in the same community, we extracted this sub-community where they are and re-detect modules of the sub-network. Specifically, we identified all nodes at the margin of the GDM community and the dementia community. Among them, we ranked and chose the union set of the top 50 quantiles of nodes in each metric (betweenness: descending; constraint: ascending) for further analysis. Note, that margin nodes (N = 18, see Methods of the supplementary file) that are ‘GDM’ or ‘dementia’ themselves or introduce noises without concrete meaning (i.e., risk reduction) were manually filtered and removed.

Next, according to the variable types from GDM to dementia, we automatically categorized all candidates into three big groups (level 1) and ten sub-groups (level 2) using GPT-4 and human validation. This study only considers the association of GDM and dementia in mothers, and thus excluded variables in fetal and birth conditions (N = 10). Further, as SemRep may imprecisely extract triples from sentences, to analyze the mediating potential of each candidate variable, we then have GPT-4 read article abstracts of each mediator candidate and score the argument quality assessing the reliability of a given mediator in connecting GDM and dementia. Please refer to the Methods of supplementary file for more methodological details of network analysis.

### LLM synthesis and reasoning on potential mechanisms

We further use GPT-4 to explore and compare potential mechanisms and clinically actionable variables based on various external knowledge bases derived from the progressive graph mining techniques above. Following the above community detection results, we designed four RAG strategies, from RAG1 to RAG4. As source sentences can provide more context information (i.e., research design) for mechanism mining, with each RAG strategy, we use research abstracts of all filtered semantic relations as an external knowledge base assisting in GDM-Dementia mechanism mining.

Specifically, RAG1 indicates the baseline GPT-4 model, with the original GPT-4 retrieval technique as the baseline results. RAG2 uses the basic GPT-4 attached with all abstracts of nature index journals as the external knowledge base. RAG3 and RAG4 are GRAG strategies. RAG3 uses the basic GPT-4 with all abstracts in the sub-community of GDM and Dementia. RAG4 uses the basic GPT-4 with all abstracts of the above Q50 intermediating variables of the subcommunity of GDM and Dementia. For each RAG strategy, we designed three questions to progressively explore the potential mechanisms, from the initial mining of potential mechanisms from GDM to Dementia, the deep mining of each generated mechanism, and the drafted plan of a long-term GDM cohort to prevent the development of Dementia based on the knowledge from last two questions. The detailed questions are demonstrated in the prompt section of the supplemental material.

### Evaluation of the GRAG process and prioritization of mediating categories

For intermediating candidates’ results and mechanism mining results, we conduct systematic evaluations separately from three perspectives: 1) the relevance and quality of the graph-based knowledge base, 2) the consistency of the RAG (GRAG)-empowered GPT-4 response and information of the refined knowledge base, and 3) the medical reliability of the aforementioned GPT-4 output from clinical expert evaluation. Considering the feasibility and reasonability of evaluations, the assessment of intermediating variables focuses on the aspects of the knowledge base and the consistency (factuality and argument quality); the evaluation of mechanism mining results concentrates on the dimensions of the knowledge base and the clinical reliability (completeness & accuracy, novelty, and clinical relevance). Note, that all scorings are scaled from 0 to 10, with 0 as not conformed at all and 10 as fully conformed.

### Knowledge base evaluation

First, a high-quality knowledge base is necessary for reliable responses. In general, we believe a knowledge base with high context precision should include up-to-date abstracts that are most relevant to the topic. The journal filtering and graph mining rules render only high-quality journal studies tightly linked to GDM and dementia eligible to the knowledge base, strictly selecting the relevant articles. Thus, we believe that both results conform to derive from a high-quality and relevant knowledge base.

### RAG response evaluation

Second, though a high-quality external knowledge base is in place, considering the hallucination in gap information of LLMs, it is crucial to check whether the GPT-4 output is consistent with the refined knowledge base. Considering the manual labor-intensive consistency evaluation, we conduct the consistency evaluation mainly for the results of intermediating variables. As for the mechanism mining results, it is not feasible to have researchers read all 30k abstracts for fact-checking and therefore we concentrate on the evaluation directly on the clinical reliability later.

Here, the consistency of the filtered knowledge base and the GPT-4 answer with its assistance is reflected by the argument quality of the response, regardless of domain knowledge not included in the filtered database or answers. We referred to the rules of evaluating an argument quality: 1) all argument elements are true (factuality); 2) they are organized logically (reasoning quality). In specific, we have two undergraduate students 1) rate all answers’ factuality by comparing all abstracts in the knowledge base and GPT-4 output based on the given knowledge; 2) rate the reasoning quality of each answer, especially logic errors like answers’ conflicts (Supplementary Table 1 for the fact-check grading criteria). Due to the generative limit (4096 tokens) of GPT-4 and the large number of variables in each mediating sub-category, GPT-4 would select several variables for evaluation rather than analyzing all candidates. After the manual evaluation, we calculate the adjusted mediating strength score for each GPT-4 chosen variable by integrating the GPT-4 mediating strength score and manual consistency assessment of the GPT-4 score. We then rank the average status of all categories of variables. Considering factuality is the foundation of high consistency in knowledge base and generation, we weigh the factuality higher than the argumentation. The equation is shown below:

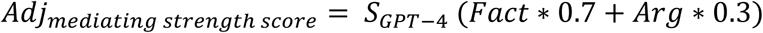

Furthermore, considering the hallucination of LLMs in evaluating large texts impacted the real status of the corresponding variable, we add one more dimension-article volume representing the evidence volume supporting the given variable candidate. Specifically, we use the proportion of abstracts of all chosen variables in a given category in all chosen variables’ abstracts as the abstract percentage of a category. When visualizing the adjusted mediating strength score and abstract percentage, the bubble size of each category is calculated below using the integrated score, *S_integrated_*. We take the natural logarithm of the abstract percentage to narrow down the difference between relatively large and small article volumes to balance the weight of evidence volume.

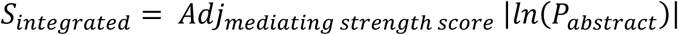

### Medical reliability evaluation

Third, we finally evaluate the medical reliability of the generated output. Regarding the strength of intermediating variables, medical reliability is assessed through existing knowledge and publications. We mainly conduct the medical reliability for the mechanism mining results via clinical expert scoring. Specifically, evaluations are conducted in three aspects, scientific accuracy & completeness, novelty, and clinical relevance. Scientific accuracy & completeness measures the degree to which the given answer complies with their domain knowledge. Novelty measures whether and the extent to which a given answer provides any content that surprises or inspires the expert. Clinical relevance assesses the feasibility and clinical relevance of the proposed tests and examinations. As all these three dimensions may influence expert scoring results, we analyzed expert scores using a single-factor analysis of variance (ANOVA). In addition, to evaluate the robustness of expert scoring, we also utilize three LLMs (GPT-4o, Llama3-70b, Gemini Adv) as LLM reviewers to score the mechanism mining results separately from these three metrics.

### UK Biobank cross-validation on identified variables and mechanisms

Finally, to validate the effectiveness of the identified variables and mechanisms, with the UK Biobank data, we measured the mediating effects of three mediator candidates of GDM-Dementia, which are *high low-density lipoprotein cholesterol (LDL-C > 3.1 mmol/L)*, *early menopause (menopause age< 40 y)*, and *depressed mood* (three representatives respectively for lipid metabolism disruptions, hormonal changes, and mental health). UK Biobank (UKB) is a tremendous population-based prospective study for investigating genetic and non-genetic factors of diseases in middle-old people [32]. Recruiting between 2006-2010 in 22 centers throughout the UK, UKB covers over a half million participants who were aged 40-69 years old since they joined and continuously collects data (the latest update in 2021) from extensive sources, such as questionnaires, physical measures, and sample assays [32]. The Ethics Committee review was approved by the North West Multi-Centre Research Ethics Committee (16/NW/0274).

This study included the same participants as our former UKB cohort study, 205,463 women participants (GDM history group: total, 1292, Dementia, 17; non-GDM history group: total, 204,171, Dementia, 2904) The covariates controlled in the mediating analysis to reduce bias effects on outcome are race (white/non-white), educational status (higher education/non-higher education), age at first live birth, multiple live births (live births count >= 5), obesity (body mass index [BMI] >= 30 kg/m^2^ at enrolment), current smoker, drinking status, physical activity, hypertension, and APOE ε4 carrier status. The mediating role of each candidate was examined using the difference method to calculate the indirect effect (GDM-mediator-Dementia) by using the overall GDM-Dementia effect minus the direct GDM-Dementia effect (controlling the mediator). The mediating proportion was the proportion of the measured indirect effect in the total effect.

## Data availability

All data generated in this study can be provided upon request. Please reach out to the corresponding authors.

## Code availability

All codes generated in this study can be provided upon request. Please reach out to the corresponding authors.

## Supplementary Materials

Supplementary Materials of this article can be found, in the online version.

## Contributions

Conceptualization: J.D., Y.Z., S.W.

Methodology: J.D., S.W., Y.Z.

Formal Analysis: S.W., D.G.

Data Curation: S.W., W.X., X.H., G.D.

Clinical Expert Evaluation: Y.Z., Y.G.

Writing—original draft: S.W., J.D., Y.Z.

Writing—review & editing: J.D., S.W., Y.Z.

Funding acquisition: J.D., Y.Z.

## Data Availability

All data produced in the present study are available upon reasonable request to the authors

## Acknowledgments

This study was funded by the National Natural Science Foundation of China (Project number 72074006 to JD), the National Key R&D Program for Young Scientists (Project number 2022YFF0712000 to JD), and the National High Level Hospital Clinical Research Funding (Interdisciplinary Research Project of Peking University First Hospital, Project number 2024IR33 to YZ, JD, and YG). We thank Ting Chen from Dublin City University of Ireland for assisting in accessing GPT-4 and other large language models for analysis.

## References

1. Wagner, C., et al., Life course epidemiology and public health. Lancet Public Health, 2024. 9(4): p. e261–e269.

2. Nakagawa, T. and G. Sala, Rethinking causal effects across the lifespan. The Lancet Healthy Longevity, 2024. 5(3): p. e170–e171.

3. Kuh, D., Y. Ben Shlomo, and S. Ezra, A Life Course Approach to Chronic Disease Epidemiology. A Life Course Approach to Chronic Disease Epidemiology. 2004. 1–494.

4. Jackisch, J. and C. Liu, Taking a life course approach to healthy ageing and multimorbidity: defining risk factors is not the end, we can do more. The Lancet Healthy Longevity, 2024. 5(1): p. e8–e9.

5. De Stavola, B.L., et al., Statistical issues in life course epidemiology. Am J Epidemiol, 2006. 163(1): p. 84–96.

6. Lewis, S.J., et al., Developing the WCRF International/University of Bristol Methodology for Identifying and Carrying Out Systematic Reviews of Mechanisms of Exposure-Cancer Associations. Cancer Epidemiol Biomarkers Prev, 2017. 26(11): p. 1667–1675.

7. Ertaylan, G., et al., A Comparative Study on the WCRF International/University of Bristol Methodology for Systematic Reviews of Mechanisms Underpinning Exposure-Cancer Associations. Cancer Epidemiol Biomarkers Prev, 2017. 26(11): p. 1583–1594.

8. Robles, L.A., et al., Does testosterone mediate the relationship between vitamin D and prostate cancer progression? A systematic review and meta-analysis. Cancer Causes Control, 2022. 33(8): p. 1025–1038.

9. Lynch, B.M., et al., Linking Physical Activity to Breast Cancer: Text Mining Results and a Protocol for Systematically Reviewing Three Potential Mechanistic Pathways. Cancer Epidemiol Biomarkers Prev, 2022. 31(1): p. 11–15.

10. Drummond, A.E., et al., Linking Physical Activity to Breast Cancer via Sex Steroid Hormones, Part 2: The Effect of Sex Steroid Hormones on Breast Cancer Risk. Cancer Epidemiol Biomarkers Prev, 2022. 31(1): p. 28–37.

11. Swain, C.T.V., et al., Linking Physical Activity to Breast Cancer via Sex Hormones, Part 1: The Effect of Physical Activity on Sex Steroid Hormones. Cancer Epidemiol Biomarkers Prev, 2022. 31(1): p. 16–27.

12. Elsworth, B., et al., MELODI: Mining Enriched Literature Objects to Derive Intermediates. Int J Epidemiol, 2018. 47(2): p. 369–79.

13. Elsworth, B. and T.R. Gaunt, MELODI Presto: a fast and agile tool to explore semantic triples derived from biomedical literature. Bioinformatics, 2021. 37(4): p. 583–585.

14. Smalheiser, N.R., et al., From knowledge discovery to knowledge creation: How can literature-based discovery accelerate progress in science*?* 2023.

15. Thilakaratne, M., K. Falkner, and T. Atapattu, A Systematic Review on Literature-based Discovery: General Overview, Methodology, & Statistical Analysis. ACM Comput. Surv., 2019. 52(6): p. Article 129.

16. Smalheiser, N.R., Literature-based discovery: Beyond the ABCs. Journal of the American Society for Information Science and Technology, 2012. 63(2): p. 218–224.

17. Peng, Y., et al., AI-generated text may have a role in evidence-based medicine. Nat Med, 2023. 29(7): p. 1593–1594.

18. Yang, J., et al., Poisoning medical knowledge using large language models. Nature Machine Intelligence, 2024.

19. Lewis, P., et al., Retrieval-Augmented Generation for Knowledge-Intensive NLP Tasks. 2020.

20. Edge, D., et al., From local to global: A graph rag approach to query-focused summarization. arXiv preprint arXiv:2404.16130, 2024.

21. Hu, Y., et al., GRAG: Graph Retrieval-Augmented Generation. arXiv preprint arXiv:2405.16506, 2024.

22. WHO. Dementia. 2023 March 15 [cited 2024 Sep 6]; Available from: https://www.who.int/news-room/fact-sheets/detail/dementia.

23. Erol, R., D. Brooker, and E. Peel, The impact of dementia on women internationally: An integrative review. Health Care Women Int, 2016. 37(12): p. 1320–1341.

24. Zhang, Y., et al., Gestational diabetes mellitus is associated with greater incidence of dementia during long-term post-partum follow-up. J Intern Med, 2024. 295(6): p. 774–784.

25. Wang, H., et al., IDF Diabetes Atlas: Estimation of Global and Regional Gestational Diabetes Mellitus Prevalence for 2021 by International Association of Diabetes in Pregnancy Study Group’s Criteria. Diabetes Res Clin Pract, 2022. 183: p. 109050.

26. Kilicoglu, H., et al., SemMedDB: a PubMed-scale repository of biomedical semantic predications. Bioinformatics, 2012. 28(23): p. 3158–60.

27. Touvron, H., et al., Llama 2: Open Foundation and Fine-Tuned Chat Models. ArXiv, 2023. **abs/**2307**.09288**.

28. Achiam, O.J., et al. GPT-4 Technical Report. 2023.

29. Kerner, S.M. GPT-4o explained: Everything you need to know. 2024 [cited 2024 October 13]; Available from: https://www.techtarget.com/whatis/feature/GPT-4o-explained-Everything-you-need-to-know.

30. Dubey, A., et al., The llama 3 herd of models. arXiv preprint arXiv:2407.21783, 2024.

31. Reid, M., et al., Gemini 1.5: Unlocking multimodal understanding across millions of tokens of context. arXiv preprint arXiv:2403.05530, 2024.

32. Sudlow, C., et al., UK biobank: an open access resource for identifying the causes of a wide range of complex diseases of middle and old age. PLoS Med, 2015. 12(3): p. e1001779.

33. Livingston, G., et al., Dementia prevention, intervention, and care: 2024 report of the Lancet standing Commission. Lancet, 2024. 404(10452): p. 572–628.

34. Christensen, M.H., et al., Kidney Disease in Women With Previous Gestational Diabetes Mellitus: A Nationwide Register-Based Cohort Study. Diabetes Care, 2024. 47(3): p. 401–408.

35. Tanaka, S. and M.D. Okusa, Crosstalk between the nervous system and the kidney. Kidney Int, 2020. 97(3): p. 466–476.

36. John, C.M., et al., Maternal Cognitive Impairment Associated with Gestational Diabetes Mellitus-A Review of Potential Contributing Mechanisms. Int J Mol Sci, 2018. 19(12).

37. Guevara-Ramírez, P., et al., Molecular pathways and nutrigenomic review of insulin resistance development in gestational diabetes mellitus. Front Nutr, 2023. 10: p. 1228703.

38. Barth, C., A. Villringer, and J. Sacher, Sex hormones affect neurotransmitters and shape the adult female brain during hormonal transition periods. Front Neurosci, 2015. 9: p. 37.

39. Knabl, J., et al., GDM Alters Expression of Placental Estrogen Receptor α in a Cell Type and Gender-Specific Manner. Reprod Sci, 2015. 22(12): p. 1488–95.

40. Real Á, D., A. Santurtún, and M. Teresa Zarrabeitia, Epigenetic related changes on air quality. Environ Res, 2021. 197: p. 111155.

41. de Luca, A., et al., Nutriepigenomics and malnutrition. Epigenomics, 2017. 9(6): p. 893–917.

42. Pan, S., et al., Unifying Large Language Models and Knowledge Graphs: A Roadmap. IEEE Transactions on Knowledge and Data Engineering, 2024. 36(7): p. 3580–3599.

43. Yao, L., et al., Exploring large language models for knowledge graph completion. arXiv preprint arXiv:2308.13916, 2023.

44. Ban, T., et al., From Query Tools to Causal Architects: Harnessing Large Language Models for Advanced Causal Discovery from Data. ArXiv, 2023. abs/2306.16902.

45. Wu, J., J. Zhu, and Y. Qi, Medical Graph RAG: Towards Safe Medical Large Language Model via Graph Retrieval-Augmented Generation. arXiv preprint arXiv:2408.04187, 2024.

46. Kresevic, S., et al., Optimization of hepatological clinical guidelines interpretation by large language models: a retrieval augmented generation-based framework. npj Digital Medicine, 2024. 7(1): p. 102.

47. *Global burden and strength of evidence for 88 risk factors in 204 countries and 811 subnational locations*, *1990-2021: a systematic analysis for the Global Burden of Disease Study 2021*. Lancet, 2024. 403(10440): p. 2162–2203.

48. Nature, S. The Nature Index journals. 2024 [cited 2024 Aug 7]; Available from: https://www.nature.com/nature-index/faq#journals.

49. NLM. SemMedDB Database Download. 2024 May 8 [cited 2024 Aug 7]; Available from: https://lhncbc.nlm.nih.gov/ii/tools/SemRep_SemMedDB_SKR/SemMedDB_download.html.

50. Blondel, V.D., et al., Fast unfolding of communities in large networks. Journal of Statistical Mechanics: Theory and Experiment, 2008. 2008(10): p. P10008.

51. Traag, V.A., L. Waltman, and N.J. van Eck, From Louvain to Leiden: guaranteeing well-connected communities. Scientific Reports, 2019. 9(1): p. 5233.

